# Relaxed peripheral tolerance drives broad *de novo* autoreactivity in severe COVID-19

**DOI:** 10.1101/2020.10.21.20216192

**Authors:** Matthew C. Woodruff, Richard P. Ramonell, Ankur Singh Saini, Natalie S. Haddad, Fabliha A. Anam, Mark E. Rudolph, Regina Bugrovsky, Jennifer Hom, Kevin S. Cashman, Doan C. Nguyen, Shuya Kyu, Michael Piazza, Christopher M. Tipton, Scott A. Jenks, F. Eun-Hyung Lee, Ignacio Sanz

## Abstract

An emerging feature of COVID-19 is the identification of autoreactivity in patients with severe disease that may contribute to disease pathology, however the origin and resolution of these responses remain unclear. Previously, we identified strong extrafollicular B cell activation as a shared immune response feature between both severe COVID-19 and patients with advanced rheumatic disease. In autoimmune settings, this pathway is associated with relaxed peripheral tolerance in the antibody secreting cell compartment and the generation of *de novo* autoreactive responses. Investigating these responses in COVID-19, we performed single-cell repertoire analysis on 7 patients with severe disease. In these patients, we identify the expansion of a low-mutation IgG1 fraction of the antibody secreting cell compartment that are not memory derived, display low levels of selective pressure, and are enriched for autoreactivity-prone *IGHV4-34* expression. Within this compartment, we identify B cell lineages that display specificity to both SARS-CoV-2 and autoantigens, including pathogenic autoantibodies against glomerular basement membrane, and describe progressive, broad, clinically relevant autoreactivity within these patients correlated with disease severity. Importantly, we identify anti-carbamylated protein responses as a common hallmark and candidate biomarker of broken peripheral tolerance in severe COVID-19. Finally, we identify the contraction of this pathway upon recovery, and re-establishment of tolerance standards coupled with a concomitant loss of acute-derived ASCs irrespective of antigen specificity. In total, this study reveals the origins, breadth, and resolution of acute-phase autoreactivity in severe COVID-19, with significant implications in both early interventions and potential treatment of patients with post-COVID sequelae.

## Introduction

In 2019, the novel betacoronavirus SARS-CoV-2 emerged from Wuhan, China, resulting in the COVID-19 pandemic^1^. With a global mortality reported around 2 percent, early clinical reporting of patients with severe disease included evidence of immune dysregulation with particular emphasis on the pro-inflammatory cytokine IL-6, invoking comparisons to so-called cytokine storms^2, 3^. These observations, alongside the early discovery of morbidity and mortality benefit of high-dose steroids in these patients were highly suggestive of an immune response in severe patients that was not only responsible for viral clearance, but may, in some cases, be contributing to overall disease pathology^4, 5^.

As immunologic studies progressed, profound alterations within the immune compartment were quickly identified as correlates of severe disease. Distinct immunotypes emerged in severe patients, including the identification of a large subset of patients that displayed increased frequencies of circulating plasmablasts, yet lacked evidence of T follicular help (Tfh)^6^. This observation was quickly bolstered by the identification of collapsed germinal center environments in patients that had succumbed to COVID-19 – a feature attributed to the overabundance of the proinflammatory mediator TNFa and the resulting failure of Tfh responses^7^. Further, a deep analysis of B cell activation pathways by our group revealed strong activation of the extrafollicular (EF) pathway – characterized by the strong induction of Tbet-driven double negative 2 (CD27^-^, IgD^-^, CD11c^+^, CD21^-^ [DN2]) B cells, large expansion of CD19^+^ antibody secreting cells (ASCs) correlated with high titers of antiviral antibodies, and depressed mutation frequencies within the ASC repertoire^8^. These findings have been confirmed in outside studies both through B cell surface phenotyping and repertoire analysis^9, 10, 11^.

A concerning feature of these B cell responses was their similarity to responses identified previously in patients with active autoimmune disease^12, 13^. In these patients, EF response activation often resulted in the *de novo* generation of new autoreactivities and correlated with disease severity^14^. At the time of our study’s publication, evidence of autoreactivity was mounting in severe disease, with observations of autoantibody-linked blood clotting^15^, anti-interferon antibodies^16^, connective tissue disease-associated interstitial lung disease (CTD-ILD)^17^, and generalized observations of clinical autoreactivity^18^. Since then, these observations have been bolstered by the reporting of broad autoreactivity within these patients – frequently targeting critical immune components including cytokine responses and linked to disease pathology in mouse modeling^19^. However, the developmental origins of these autoreactivities, their connection with the underlying *de novo* antiviral response, and their ultimate resolution remain unknown.

Here, using single-cell B cell repertoire analysis in highly characterized patients with severe disease, we identify a unique compartment of ASCs, primarily identified by low mutation frequencies and preference for IgG1 class switching. These cells display relaxed selective pressures and incorporate increased levels of B cell receptor configurations associated with generalized autoreactivity. While highly enriched for antiviral targeting, we identify clonotypes that are simultaneously viral antigen-responsive and autoreactive against naive B cells. Emergence of this population is correlated with clinical autoreactivity, and through broad clinical autoreactivity screening, we identify an unexpected biomarker of the generalized relaxation – anti-carbamylated protein responses. These antibodies have clear pathological potential, with a subset of patients displaying antibodies against glomerular basement membrane which are frequently associated with kidney and lung injury. Finally, we track these responses over the course of patient recovery and identify the contraction and ultimate resolution of the acute-phase autoreactive ASC compartment.

## Results

### Extrafollicular B cell-correlated ASCs lack surface receptor expression

Previous work has established the robust expansion of the ASC compartment as a hallmark of severe COVID-19. Retrospective analysis of previously collected data from 25 (HD = 9; OUT-C = 7; ICU-C = 9) patients revealed an unexpected expansion of CD19^-^ ASCs, further confirming the prominence of ASC expansion as a significant fraction of the overall acute B cell response in severe/critical COVID-19 (Fig 1a,b, Supplemental Figure 1, Supplemental tables 1,2). ASC expansion in the ICU-C cohort was directly correlated with expansion of DN2 B cells (CD19^+^, IgD^-^, CD27^-^, CD21^-^, CD11c^+^), an important intermediate in the extrafollicular (EF) B cell response pathway previously identified in both rheumatic disease and severe COVID-19 (Fig 1c)^8, 12^. Indeed, by calculating the ratio of EF (DN2 and activated naive) vs. germinal center (DN1, switch memory) intermediates in these patients, the emphasis of the EF pathway in the ICU-C cohort could be readily identified (Fig 1d).

**Figure 1.**
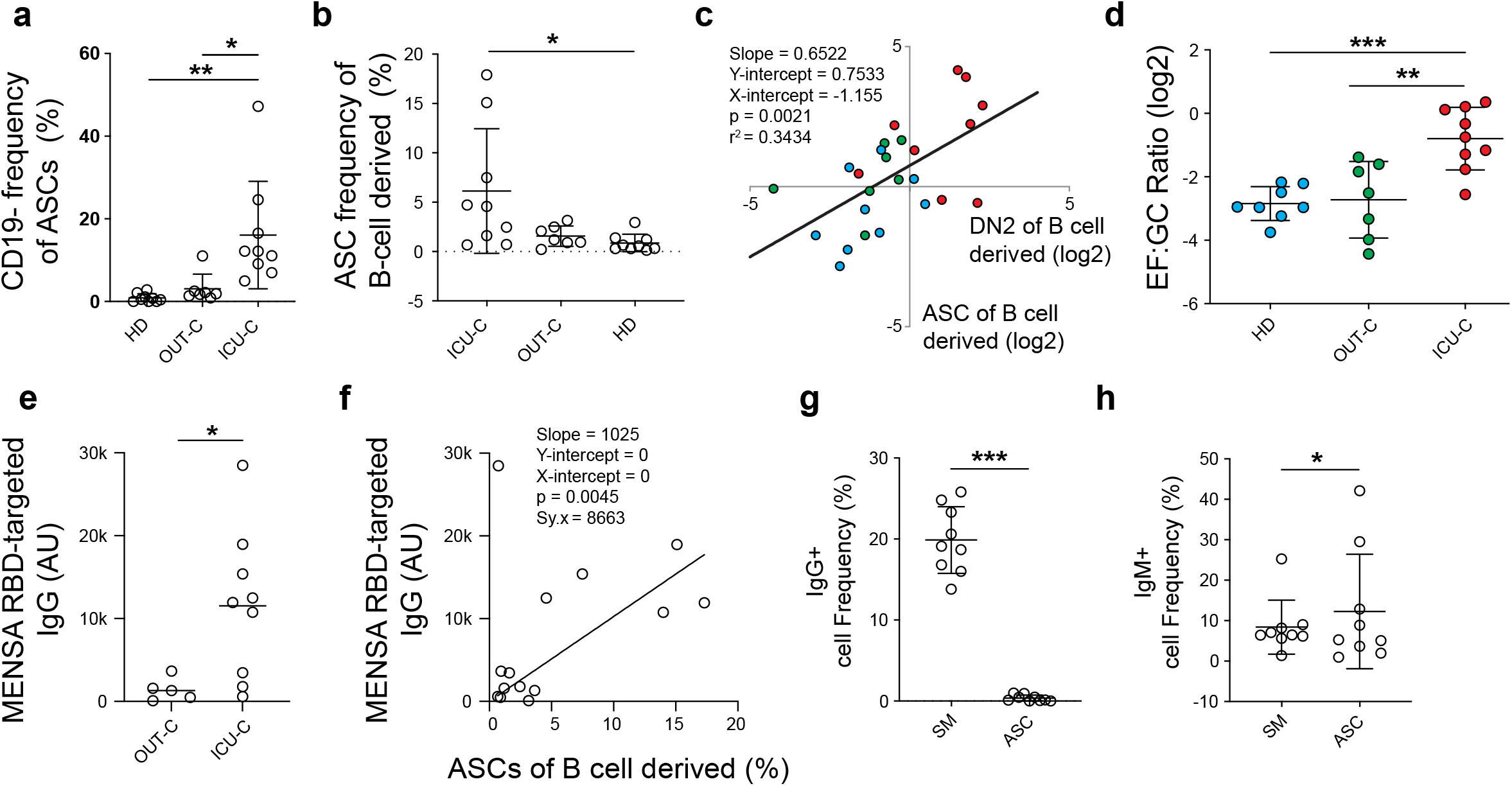
Extrafollicular B cell-correlated ASCs lack surface receptor expression **(a-d, f-g)** PBMCs from HD (n = 9), OUT-C (n = 7) or ICU-C (n = 9) patients were analyzed by FCM. (**a)** CD19^-^ ASC frequency of total ASCs. (**b)** ASC frequency of total B cell-derived cells. (**c)** Linear regression analysis of log2-transformed DN2 vs ASC frequencies of total B cell-derived cells. **(d)** Log2 expression of EF:GC B cell ratio (see methods). (**e)** RBD-specific IgG antibody in MENSA samples collected from OUT-C and ICU-C patients. (**f)** Linear correlation of RBD-specific IgG antibody in MENSA samples vs. ASC frequency of B cell-derived cells in OUT-C and ICU-C patients. (**g)** IgM^+^ frequency of total SM or ASC populations. (**h)** IgG^+^ frequency of total SM or ASC populations. Statistical significance was determined using ANOVA with Tukey’s multiple-comparisons testing between all groups. *P ≤ 0.05; **P ≤ 0.01; ***P ≤ 0.001; ****P ≤ 0.0001. Summary statistics (c–k): mean ± standard deviation (s.d.).

Previous work has correlated expansion of the ASC compartment correlated with increased serological IgG responses to the SARS-CoV-2 spike protein receptor binding domain (RBD) in patients with severe disease, however it did not directly test antigen specificity^8^. Using a novel in vitro culture method, ASCs from the ICU-C and OUT-C cohorts were cultured overnight and the supernatant, or media enriched in newly synthesized antibodies (MENSA) was collected^20^. MENSA analysis, which is more specific to the circulating ASC compartment, revealed an increased cellular component in the blood of these ICU-C patients capable of secreting IgG RBD-specific antibodies, and confirming the relevance of early circulating ASCs to antiviral response as opposed to non-specific cellular expansion (Fig 1e). Indeed, overall IgG-switched RBD-targeted MENSA titers were directly correlated with ASC expansion across the COVID-19^+^ cohorts (Fig 1f).

As the EF response pathway and ASC expansion have been previously linked with the detection of autoreactive antibodies in the serum, and these pathogenic antibodies have now been linked to COVID-19, it was important to understand the ASC contribution both to antiviral and autoantigen targeting. However, direct study of these cells is difficult due to their propensity to downregulate surface B cell receptor (BCR). Despite previous identification of expanded IgG ASC compartments in the repertoire, and the confirmation of IgG RBD-directed ASCs in MENSA assessment, IgG was almost completely absent from the surface of the ASC compartment in contrast to the switched memory (Fig 1g). Of interest, IgM was retained on the surface of ASCs, potentially suggesting a continuing signaling requirement in these cells (Fig 1h). In total, these findings suggest that due to inconsistent BCR surface expression, antigen-specific flow-based studies are likely to result in overestimation of the IgM ASC contribution to the overall antigen-specific response. As a result, broad analysis of this cellular compartment independent of BCR expression and antigen-specific probing was required.

### Expansion of low-selection IgG1 ASC compartment is a hallmark of severe COVID-19

To comprehensively study the nature of the ASC compartment within these patients, 7 of 10 recruited ICU patients, alongside 3 demographically matched healthy donors were selected for single cell VDJ repertoire (scVDJ) analysis. In total, more than 20,000 ASCs from 14,000 clonotypes were assessed (Supplemental table 3).

A broad assessment of ASCs compartment across the cohort confirms previous identification of an oligoclonal expansion of ASCs, with data from several patients reflecting more than 500 independent ASC clonotypes (Supplemental table 3). Assessment of ASC isotype across the cohort was remarkably consistent, with IgG1^+^ ASCs making up a significant fraction of ASCs in the ICU-C cohort in comparison to the IgA-dominated healthy donor ASC compartments (Fig 2a). This expansion of IgG1^+^ ASCs was statistically significant and came at the expense of slight decreases in IgG2, IgA1, and IgA2 ASC members (Fig 2b). Identification of these IgG1 clonotypes was consistent with both bulk serology, where total IgG1 was elevated in the ICU-C cohort (Supplemental figure 2a). In addition, these findings were consistent with a retrospective analysis of published single cell transcriptomics data collected from bronchoalveolar lavage fluid (BALF) of 10 intubated patients which identified substantial IgG1 expression in the plasmablast population (Supplemental figure 2b)^21^.

**Figure 2.**
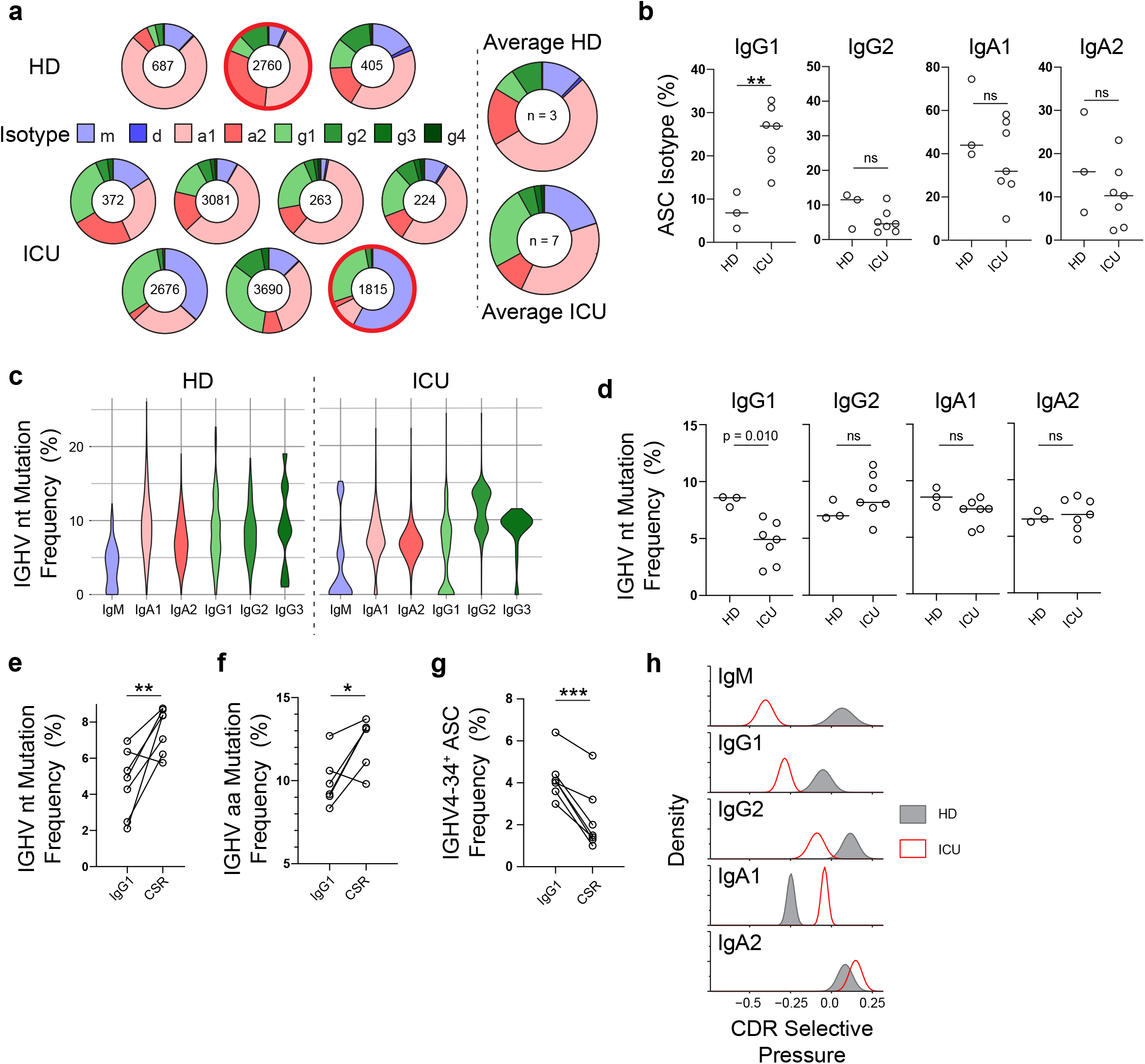
Expansion of low-selection IgG1 ASC compartment is a hallmark of severe COVID-19 **(a-h)** Single cell VDJ analysis of ASCs from HD (n = 3) and ICU-C (n = 7) cohorts **(a)** Left -individual ASC isotype compositions of HD ICU-C patients; right – averaged ASC isotype compositions by group of ICU-C patient samples. **(b)** Frequency of indicated ASC isotypes of total ASCs. **(c)** Representative ASC mutation frequency distributions by isotype in HD, vs ICU-C patient. **(d)** IGHV-gene nucleotide mutation frequencies of indicated ASC isotypes in HD or ICU-C patients. **(e)** IGHV-gene nucleotide mutation frequencies of IgG1, versus other class-switched ASCs. **(f)** IGHV-gene amino acid mutation frequencies of IgG1, versus other class-switched ASCs. **(g)** IGHV4-34^+^ ASC frequency in IgG1, versus other class-switched ASCs. **(h)** BASELINe selection analysis of CDR selection in HD, versus ICU-C ASCs, grouped by isotype.

Focusing in on this expanded compartment, mutation analysis of the IgG1^+^ ASCs in patients across the cohort displayed a dramatic reduction in mutation frequency in comparison to HD (fig 2c,d). This reduction in mutation frequency was largely specific to the IgG1 compartment with IgG1 ASC mutations frequencies significantly decreased in comparison to the rest of the class-switched ASC compartment (Fig 2e,f). Although some patients displayed broader suppression of mutation frequencies across multiple isotypes^8^, this phenotype was restricted to a fraction of the overall cohort (fig 2d).

Previous studies in SLE have identified the emergence of autoreactive B cell clonotypes concomitant with reduced mutation frequencies in the ASC compartment due to a relaxation of peripheral tolerance^14^. An important bellwether of this phenomenon is the expansion of B cell clonotypes containing the autoreactivity-prone V gene *IGHV4-34*. Concerningly, these trends were reflected in the repertoire of the ICU cohort with increased frequency of IGHV4-34 positive cells emerging specifically within the IgG1^+^ ASC compartment (Fig 2g). Consistent with these observations, an overall analysis of the selective pressure on the antibody complementarity determining regions, as determined by Bayesian estimates of antigen-driven selection (BASELINe)^22^, revealed a selective reduction in the IgG1, but not other class-switched compartments in comparison to ASCs from healthy donors (Fig 2h). Overall, these data reflect an expanded, low mutation compartment within the overall ASC response marked by low selective pressures and a predilection towards autoreactivity-prone BCR incorporation.

### IgG1 ASCs display evidence of *de novo* generation

To more deeply understand the origins and persistence of the low-mutation IgG1 ASC compartment, the B cell memory, and CD27-(non-memory) compartments were additionally sorted and analyzed in 4 surviving patients from the original ICU cohort (Supplementary table 1). Of the patients selected for further study, two displayed sufficient cellularity and lineage diversity in the IgG1^+^ ASC compartment to justify connectivity analysis in assessing cellular origins.

Interestingly, while both patients displayed reduced mutation frequency distributions within the IgG1 compartment, these reductions were not identifiable in the contemporaneously collected memory (Fig 3a). This correlated with normal levels of selective pressure within the memory compartment suggesting a lack of penetration of low selection clonotypes (Fig 3b). An important reflection of the resistance of memory to clonotypes emerging under relaxed selective pressures was identified in the autoreactivity-prone *IGHV4-34* expressing cells. In germline configuration, these cells often display self-reactivity against a variety of autoantigens mediated through the antibody framework region 1 (FR1), and independent of complementarity-determining regions (CDRs). These interactions are frequently dependent on the presence of an amino acid ‘patch’ encoding the hydrophobic amino acid sequence A-V-Y in FR1. Under normal selective pressure, these cells are routinely selected against, and their incorporation into activated B cell compartments is frequently coupled with the disruption of the AVY patch through somatic hypermutation^23^. Importantly, while *IGHV4-34*^+^ ASCs in these ICU patients retained the AVY patch with more than 90% frequency, contemporaneous memory continued to display evidence of AVY-censoring (Fig 3c).

**Figure 3.**
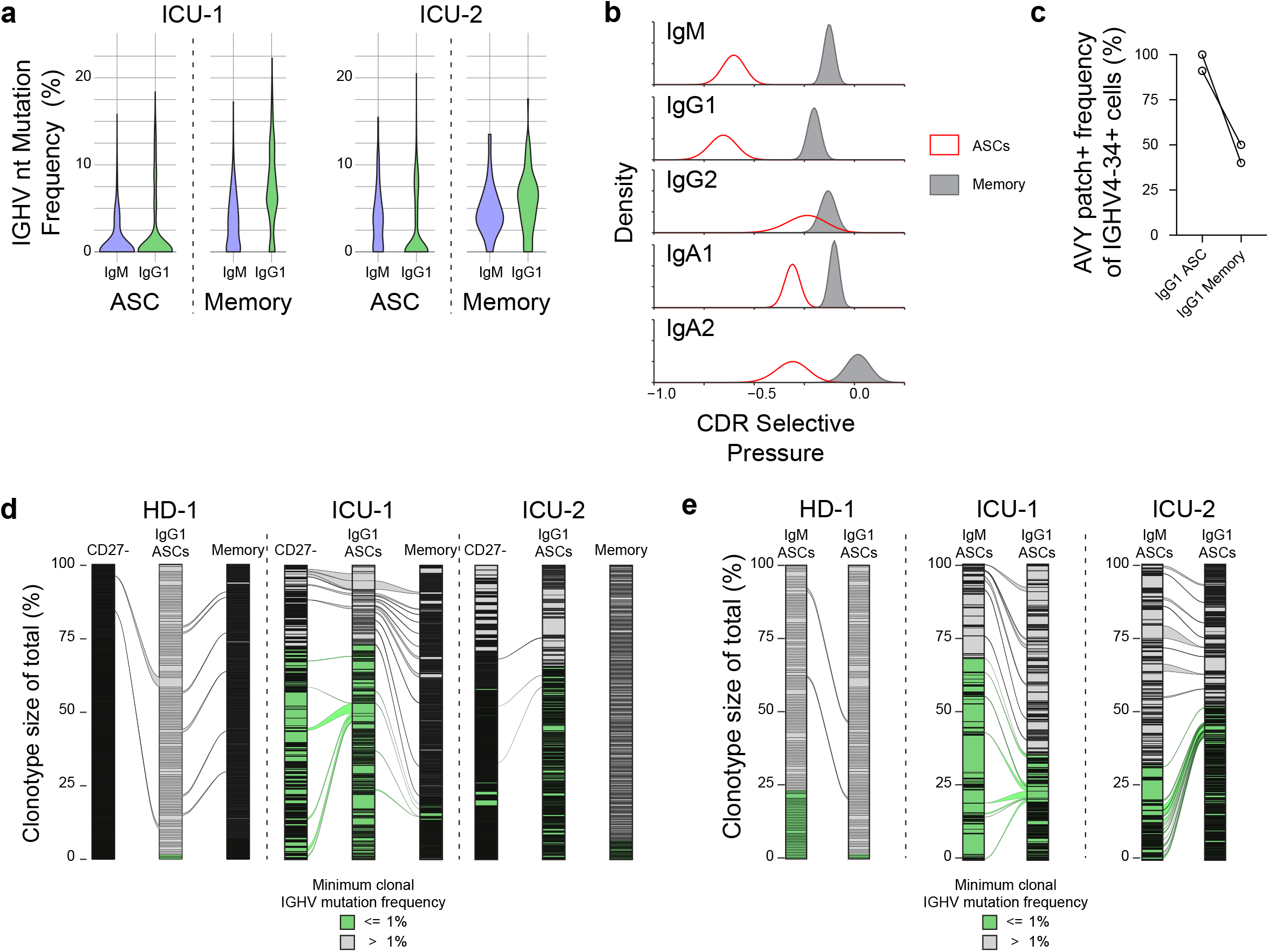
IgG1 ASCs display evidence of de novo generation **(a-h)** Single cell VDJ analysis of ASCs, memory, and CD27-compartments from ICU-C patients **(a)** IGHV nucleotide mutation frequency distributions by isotype in ASC vs. memory B cell populations (n = 2). **(b)** BASELINe selection analysis of CDR selection in ASC vs. memory B cell populations, grouped by isotype (n = 4). **(c)** AVY^+^ ASC frequency of total IGHV4-34^+^ ASC frequency in IgG1 ASC versus IgG1^+^ memory B cell populations. **(d)** Alluvial plots showing clonotype connectivity between IgG1 ASCs to the CD27^-^ or memory compartments. Individual clonotypes represented by vertical banding, with the height of the band reflective by the number of cells incorporated into the clonotype. Clonotypes with minimum mutation frequencies <= 1% are highlighted in green. **(e)** Alluvial plots showing clonotype connectivity between IgG1 ASCs to the IgM ASC compartment. Clonotypes with minimum mutation frequencies <= 1% are highlighted in green.

These findings suggested that rather than being memory-derived, the IgG1 ASCs represent an independently derived compartment that might instead be connected to the naive or double-negative compartment expansions previously identified in severe COVID-19. Direct visualization of shared clonotypes between low-mutation IgG1 ASCs and the surrounding B cell response showed a preference for connections with the CD27-compartment, particularly in the largest expanding clones (Fig 3d). Indeed, patient ICU-2 lacked *any* connection with the memory compartment in the acute phase of disease. In addition to a lack of connection to the surrounding memory compartment, low-mutation (and germline) IgG1 ASCs were highly connected to IgM ASC counterparts suggesting an ongoing class-switch event within the clonotype at the time of PBMC isolation (Fig 3e). Altogether, these data strongly suggest that this unique compartment is polyclonal, mostly derived independently from memory, and is instead rooted in an IgM^+^ CD27^-^ B cell compartment – all highly suggestive of a rapidly-developing naive-derived response.

### IgG1 clonotypes are both anti-viral and autoreactive

As previous studies of antigen-specific responses have likely underrepresented analysis of this unique IgG1 ASC compartment due to a lack of surface receptor expression (Fig 1g), it was important to understand the specificity of these clonotypes and understand their relevance to the overall antiviral responses previously documented in the literature. To this end, 55 clonotypes were selected from patient ICU-1 for monoclonal antibody production and screening (Supplemental table 4). Clonotypes were selected based on their inclusion of an IgG1 member, low minimum mutation frequency (<1%), and presence in the ASC compartment, CD27-compartment, or both (Fig 4a). In addition to all expanded clonotypes (> 5 members) that met these criteria, all *IGHV4-34*-expressing members were included in the screening analysis.

**Figure 4.**
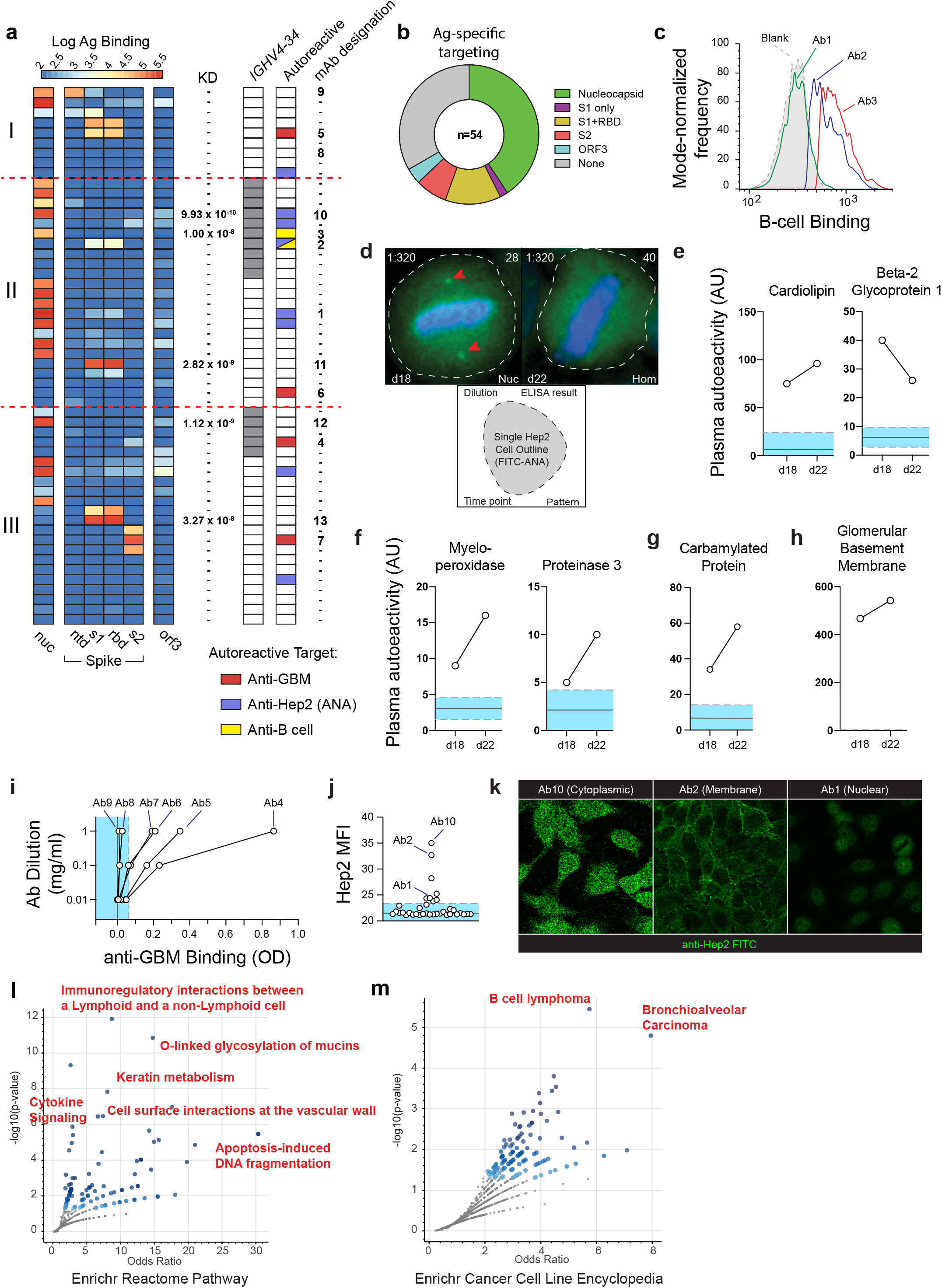
IgG1 clonotypes are both anti-viral and autoreactive (a) Overview of mAb testing from patient ICU-1. Clonotypes were selected from the CD27-compartment (III), ASCs (II), or both (I). Left -heatmap of mAb (rows) binding to indicated antigens (columns). Middle – antibody affinities confirmed through HT-SPR. Right – IGHV4-34 status and identified autoreactivities. (b) SARS-CoV-2 antigen targeting across all 55 mAbs. (c) Naive B cell binding of two monoclonal antibodies as identified in (a). (d) Immunofluorescent assessment of anti-nuclear antibodies in plasma from patient ICU-1 at indicated time points via Exagen-performed clinical testing. (e-g) ELISA-based assessment of indicated autoantigens in plasma from patient ICU-1 at indicated time points via Exagen-performed clinical testing. Blue bars represent 3x standard deviation of healthy donor control plasma. (e) Antiphospholipid syndrome-associated autoantigen assessment of ICU-1 plasma. (f) Autoimmune vasculitis-associated autoantigen assessment of ICU-1 plasma. (g) Anti-carbamylated protein antibody assessment of ICU-1 plasma. (h) Anti-glomerular basement membrane (GBM) antibody assessment of ICU-1 plasma. (i) Anti-GBM ELISA testing of isolated monoclonal antibodies. Select mAb designations indicated [Fig 4a] (j) Mean fluorescence intensity (MFI) measurements of Hep2 cell line reactivity by synthesized monoclonals via immunofluorescence. Select mAb designations indicated [Fig 4a,k] (k) Representative staining patterns from select mAbs with reactivity against the Hep2 cell line. (l) Reactome pathway analysis of z-score-ranked list of autoreactivities identified by CDI autoantigen proteomics. (m) Cell cancer line encyclopedia enrichment analysis of z-score-ranked list of autoreactivities identified by CDI autoantigen proteomics.

Using a luminex-based approach^24^, mAbs were screened against multiple SARS-CoV-2 antigens including S1, RBD, NTD, S2, ORF-3, and nucleocapsid (Fig 4a). Of the 55 Abs tested, more than 70% showed affinity to one of the tested target antigens, with a surprising frequency of nucleocapsid-targeted responses (Fig 4b). Despite their naive origin, low somatic hypermutation, and reduced selective pressures, many of the resulting antibodies were of reasonably high affinity with KDs in the low nanomolar range, with the top binders to the spike and nucleocapsid proteins displaying affinities of 2.82 ⨯ 10^−9^ and 9.93 ⨯ 10^−10^, respectively (Fig 4a, Supplemental Figure 3). Of interest, *IGHV4-34*-expressing clones were generally viral-targeted, with most targeting the nucleocapsid protein (Fig 4a). Overall, these data strongly suggest that this compartment is highly enriched for antigen-specific ASCs and relevant to the emerging antiviral response.

However, despite the presence of high levels of antigen specificity, 30% of the clones tested did not show clear specificity to any viral protein, and many displayed low binding activities (Fig 4a). Due to the emerging literature surrounding autoreactivity in COVID-19, and the low selective pressures displayed across this compartment, it was important to understand if any of these antibodies also contained autoreactive potential. To test this directly, monoclonals were screened for the capacity to bind human naive B cells – a feature of some *IGHV4-34* clones previously identified in systemic lupus erythematosis (SLE) linked to reactivity against I/i antigen on the B cell surface^23^. Concerningly, 2 of the 55 antibodies displayed positive binding activity despite displaying binding potential for SARS-CoV-2 antigens confirmed through surface plasmon resonance (Fig 4a,c, Supplemental figure 3). Importantly, one of the clones displayed reactivity against the SARS-CoV-2 receptor binding domain (RBD), confirming the naive derivation of autoreactivity in severe COVID-19 as RBD responses are highly specific to SARS-CoV-2^25^.

As this autoreactivity-prone IgG1 compartment is clearly expanding within the ongoing response, it was relevant to understand if clinical autoreactivity levels were building over the acute phase of infection. To test this, plasma was collected from patient ICU-1 at d18, and d22 post symptom onset, and tested for clinical autoreactivity by Exagen’s clinical laboratory. Of concern, this patient displayed elevated test results to 7 independent clinically relevant antigens of which 6 were actively building over the 4-day time period between draws (Fig 4d-f). In addition to the autoreactivities that have been previously associated with severe COVID-19 including anti-nuclear antibodies (ANAs) and antibodies associated with antiphospholipid syndrome (Fig 4d,e). These activities were accompanied by indicators of autoimmune vasculitis with reactivities identified against both myeloperoxidase and proteinase 3 (Fig 4f). Unexpectedly, we also identified a high level of anti-carbamylated protein antibodies which has been previously linked to disease activity in both SLE and RA (Fig 4g). Perhaps most concerningly, this patient displayed remarkable levels of anti-glomerular basement membrane antibodies directly implicated in glomerular destruction in the kidney and alveolar destruction in the lung in extremely rare patients with Goodpasture’s syndrome (Fig 4h)^26^. These identified plasma autoreactivities were directly linked to the low-selection ASCs underpinning this response through the identification anti-GBM reactivity of varying affinity in 4 of the 55 synthesized mAbs (Fig 4a, i). Of particular interest was mAb5, which displayed intermediate affinity to both RBD and GBM, again confirming the naive-derived nature of these autoreactive responses. Again, in accordance with the patients autoreactivity profile, an additional 8 mAbs were identified as reactive against the Hep2 cell line commonly used for ANA-based testing (Fig 4j). Of great interest was the diversity of specific antigen targeting even within ANA clinical testing, with specific antibodies targeting independent antigens resulting in cytoplasmic, membrane, and nuclear staining of the Hep2 line (Fig 4k).

The clinical findings, antibody specificities, and relaxed selective pressures identified within the emerging ASC compartment were strongly suggestive of a broad relaxation of peripheral tolerance leading to scattered autoreactivity as has been recently described against immune components^19^. Using a proteomics-based approach, we identified a similar breadth of autoreactivity, resulting in more than 250 significant individual hits in this patient including autoreactive targeting of IFNa, CD69, L-selectin, and SLAMF7 (Supplemental table 5). Pathway analysis of these cumulative hits were highly suggestive that these breaks in tolerance were not random and were instead directed against components of the local milieu with autoreactivities against cytokine signaling, apoptotic DNA fragmentation, and immunoregulatory interaction pathways all displaying significant enrichment within this patient (Fig 4l, Supplemental figure 4). Consistent with this finding, the autoreactive profiles of this patient significantly reflected the gene expression profiles of two previously characterized cancer cell lines derived from B cell lymphoma and bronchoalveolar carcinoma (Fig 4m). Altogether, these data strongly suggest that the relaxed selective pressures and autoreactive tendencies of the low-mutation IgG1 repertoire reflect a broad relaxation of peripheral tolerance whereby autoreactivities are generated against a variety of antigens readily available in the milieu of severe acute lung infection.

### Characterizing clinical autoreactivity profiles in COVID-19

To understand the breadth and clinical relevance of this relaxation of peripheral tolerance, plasma collected from 27 ICU-C, 18 OUT-C, 20 SLE, and 14 HD were retrospectively assessed through clinical testing at Exagen and analyzed for generalized autoreactivity across the cohort. In alignment with the patient–specific data above, broad tolerance breaks were identified across the ICU cohort against a variety of targets including rheumatoid factor, phospholipids, nuclear antigens, and glomerular basement membrane (Table 1). While the majority of ICU patients displayed at least one clinically identifiable autoreactivity, some patients were much higher with one patient testing positive for 7 independent autoantigens (Fig 5a). This trend towards higher ‘densities’ of autoreactivity was significantly increased in ICU-C patients over both HD, and OUT-C cohorts (Fig 5a).

**Figure 5.**
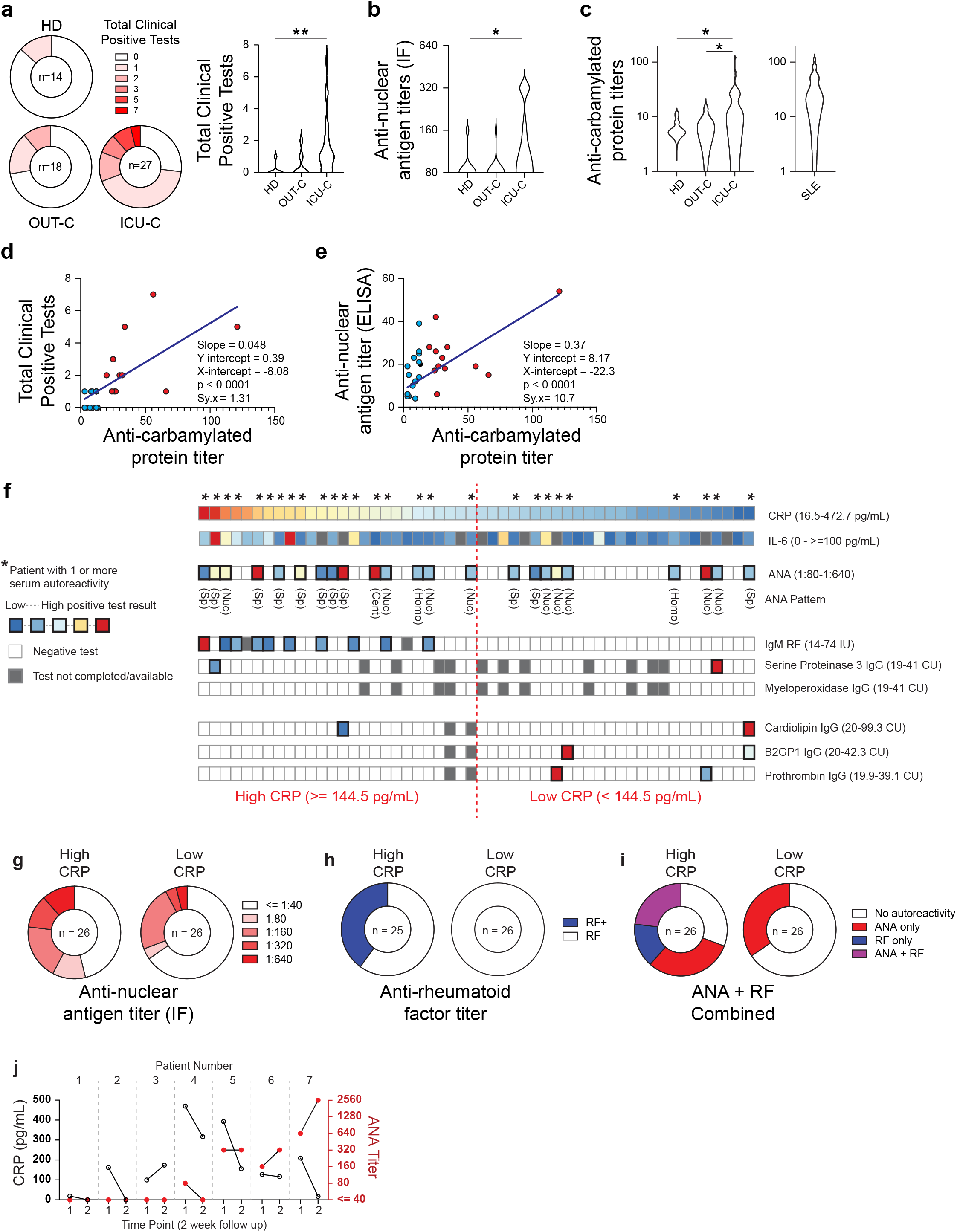
Characterizing clinical autoreactivity profiles in COVID-19 (a-d) HD, ICU-C, OUT-C, and active SLE patient frozen plasma was tested against a variety of autoantigens in Exagen’s clinical laboratory. (a) Frequency (left) and distribution (right) of total positive clinical tests across the HD, OUT-C, and ICU-C cohorts. (b) Distribution of anti-nuclear antigen titers across the HD, OUT-C, and ICU-C cohorts. (c) Distribution of anti-CarP titers across the HD, OUT-C, and ICU-C cohorts (left), and active SLE cohort (right). (d) Linear regression of anti-carbamylated protein titers vs. total number of patient autoreactive breaks across the ICU-C cohort. Patients with positive anti-CarP titers are highlighted in red. (e) Linear regression of anti-CarP protein vs. ANA titers across the ICU-C cohort. Patients with positive anti-CarP titers are highlighted in red. (f) Heatmap display of Emory pathology-confirmed clinical results of 52 SARS-CoV-2 ICU patients with US NIH “severe” or “critical” clinical designations. Patients are organized by ascending CRP values (range 16.5-472.7). Individual testing scale values are indicated following the test name. (g) Frequency of anti-nuclear antigen titers in high vs. low CRP patients within the independent ICU cohort. (h) Frequency of rheumatoid factor positive tests in high vs. low CRP patients within the independent ICU cohort. (i) Frequency of anti-nuclear antigen and rheumatoid factor positive tests in high vs. low CRP patients within the independent ICU cohort. (j) 2-week follow up testing of 7 patients from the independent ICU cohort. C-reactive protein and anti-nuclear antigen titers are displayed.

Broad clinical testing of the ICU-C cohort identified the statistically significant emergence of two autoreactivities – anti-nuclear antigen (ANA) and anti-carbamylated protein responses (CarP) (Fig 5b,c). While ANAs are well characterized in the literature in their association with autoimmunity, interpreting results can be clinically challenging as significant numbers of healthy patients with no rheumatologic symptoms can test positive in immunofluorescence-based assays. Nonetheless, more than 40% of the ICU-C cohort displayed reactivities at titers higher than 1:160 – significantly higher than expected in a randomly assessed healthy population^27^. Anti-CarP antibodies, however, were highly specific to the ICU-C cohort with more than 40% displaying positive clinical tests (Table 1, Fig 5c). Levels of anti-CarP antibodies were high, with OUT-C patients displaying similar titers to active SLE patients tested alongside the cohort (Fig 5c). Of great interest, titers of a-CarP were directly correlated with the overall number of tolerance breaks across the cohort (Fig 5d). Their further correlation with ANA titers and high specificity to the ICU-C cohort suggest that anti-CarP antibodies could serve as an important biomarker of acute peripheral tolerance breaks in severe COVID-19 if widely implemented (Fig 5e, Table 1).

Due to the strong activation of the EF pathway in the ICU-C cohort, and our previous descriptions of the pathway in active SLE, an interesting aspect of the autoreactivity data was the lack of positive testing for ‘reflex’ nuclear antigens such as Jo, Sm, La, and Ro, as well as the canonical SLE autoantigen, dsDNA (Table 1). Again, this suggests that breaks in tolerance are not random in nature, and instead likely reflect the availability of specific autoantigens within the broader inflammatory milieu.

While the initial cohort was tested using banked frozen plasma, it was important to understand the correlation of readily testable autoantigens with disease severity through fresh clinical testing in the ICU. To this end, a retrospective observational study was conducted including 52 independent critically ill patients who were admitted to COVID ICUs in Atlanta, Georgia to assess for correlations between autoantibodies and disease severity (Fig 5f). Patients in this cohort were included if they had received autoantibody testing as part of routine clinical care at the discretion of their treating physicians. Validating our banked plasma cohort, more than 50% of patients in the fresh collection cohort tested positive for at least one autoreactivity with ANAs as the most common autoreactive feature (a-CarP antibody testing was not available) (Fig 5g). Once again, disease severity directly correlated with tolerance breaks as patients displaying the highest C-reactive protein levels (CRP – a surrogate of disease severity in COVID-19^28^) displayed both increased numbers and intensities of autoreactive tests (Fig 5f-i). While follow up testing for this cohort was limited in availability, 7 patients were tested 2 weeks after the initial draw with 3 of 7 testing positive for ANAs on initial assessment (Fig 5j). Of concern, but in alignment with the building autoreactivity pattern identified in patient ICU-1, all three patients displayed stable or increasing ANA titers over the two-week period despite decreased CRP, suggesting a building autoreactivity profile in these patients beyond the resolution of other biomarkers of clinical illness.

### Relaxed peripheral tolerance largely resolves upon recovery

With autoreactivity profiles building in acute disease in ICU-C patients (Fig 4h-j, 5h-j), it was important to understand the persistence of the permissive IgG1 ASC repertoire and associated autoreactivities in the recovery phase of disease. Reassessment of patient ICU-1 6 months following recovery revealed a drop in autoantibody levels against many of the antigens identified in the acute phase of disease to healthy donor-levels – including anti-GBM (Fig 6a). Of great interest, concomitant with this drop was a loss of *all* of the B cell clonotypes that were selected for mAb production and testing from the recovery-phase repertoire. An analysis of connectivity between the acute ASC, and recovery memory compartment revealed that while most isotypes displayed conversion into memory, the IgG1 fraction showed little persistence (Fig 6b). This loss was reflected in contraction of the IgG1 ASC compartment in patients that had displayed robust expansion, with an associated expansion in IgG2 (Fig 6c).

**Figure 6.**
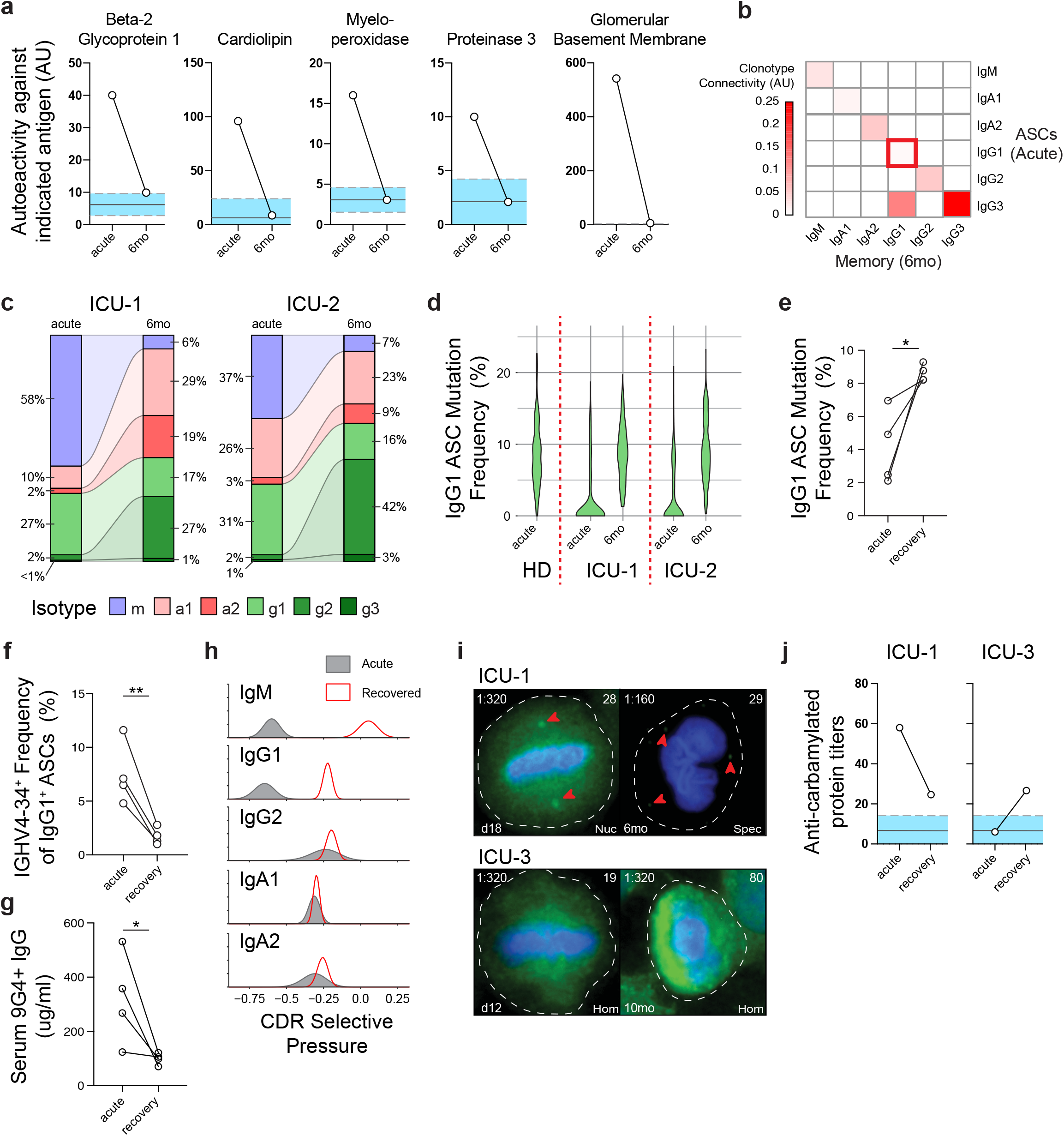
Relaxed peripheral tolerance largely resolves upon recovery. (a) Exagen clinical testing results of patient ICU-1 plasma during the acute phase and 6 months post recovery. Blue bars represent 3x standard deviation of healthy donor control plasma. (b) Clonotype connectivity heatmap displaying strength of longitudinal connections between acute-phase ASCs and 6 month recovery memory B cells in patient ICU-1. Connectivity values represent the Simpson’s Index of beta diversity across populations subtracted from 1. (c) Isotype frequencies of ASC populations in patients ICU-1 and ICU-2 at acute and recovery time points (6mo). (d) *IGHV* nucleotide mutation frequency distributions in IgG1 ASCs in HD, ICU-C acute, or ICU-C recovery samples. (e) *IGHV* nucleotide mutation frequency in IgG1 ASCs in acute vs recovery samples in all 4 ICU-C single-cell patients. (f) *IGHV4-34*^*+*^ ASC frequency in IgG1 ASCs in acute vs recovery samples in all 4 ICU-C single-cell patients. (f) ELISA assessment of *IGHV4-34*^*+*^ IgG plasma antibody concentration. (h) BASELINe selection analysis of CDR selection in ASCs at acute vs. recovery timepoints of all 4 ICU-C single-cell patients, grouped by isotype. (i) Immunofluorescent assessment of anti-nuclear antibodies in plasma from patients ICU-1 and ICU-3 at indicated acute and recovery time points. (j) Anti-carbamylated protein antibody assessment of ICU-1 plasma through Exagen clinical laboratory testing.

Importantly, in all 4 patients that survived to the recovery time point, the low-mutation phenotype identified in acute disease within the IgG1 compartment had resolved to reflect a ‘normal’ resting HD mutation distribution (Fig 6d,e). This increase in mutation frequency was accompanied by a reduction in the inclusion of *IGHV4-34* clones within the IgG1 compartment, and a decrease in total *IGHV4-34+* IgG antibodies in the plasma, as assessed by the anti-idiotype antibody 9G4 (Fig 6f,g). Importantly, the IgG1 isotype compartment displayed a return to ‘normal’ selective pressure reflective of the other class-switched ASC compartments upon recovery (Fig 6h). However, despite these positive signs of a return to ‘normal’ tolerance environment within the ASC compartment, select autoreactivities, particularly ANAs and anti-CarP responses, persisted or even progressed over the recovery period, demanding continued study into other potential reservoirs of newly-generated autoimmune memory (Fig 6i,j).

## Discussion

While several studies surrounding autoreactivity in COVID-19 have detailed the presence of autoantibodies in COVID-19, their connections to the underlying immune repertoire, cellular origins and kinetics, and eventual resolution have remained unclear. Here, we describe a unique compartment of IgG1-class switched ASCs that emerge as a major component of the early humoral immune response to infection. These cells are not memory derived – displaying active connections to CD27^-^ compartments and non-class switched ASCs and having undergone little, if any, somatic hypermutation. They display reduced selective pressures, and an associated increase in incorporation of the germline-autoreactive V gene *IGHV4-34*.

Although largely targeted against viral proteins, these low-selection cells can produce antibodies that are simultaneously specific for SARS-CoV-2 (even at moderate affinity), and also self-antigen. The nature of this self-antigen is important – while the ability to bind naive B cells has been described previously in conjunction with autoimmune disease, in COVID-19 this observation is coupled with broad targeting of the immune compartment including, in our proteomics-based dataset, BCR light chain. Indeed, the breadth of autoreactive targeting was significantly enriched for genes overexpressed in primary B cell lymphoma suggestive of a targeted break of tolerance against immune activation in the local milieu. This interpretation of the data is additionally supported by the work of Yang et. al. identifying inflammatory cytokines --many linked to autoreactive B cell responses --as a primary target of broad autoreactivity. These findings, alongside the *de novo* nature of the resulting responses, are highly suggestive of a robust developmental B cell activation environment where relaxed peripheral tolerance results in mixed targeting of viral and self-antigen – particularly self-antigen present in the surrounding EF developmental niche.

Further evidence for this model of acute relaxed peripheral tolerance can be identified in the targeting of lung tissue by the autoreactive response. While broad tolerance breaks were identified most significantly with genes expressed in B cell lymphoma, the highest strength of association was identified in bronchoalveolar carcinoma – a cancer specific to the lung epithelia <u>without</u> invasion into the lung parenchyma. Restated, patient ICU-1 displayed broad autoreactivity against genes specifically overexpressed in the primary cellular targets of SARS-CoV-2 in the lung. This, alongside the identification of rare anti-GBM antibodies frequently associated with alveolar pathology in more than 10% of our ICU cohort, and previous reporting of lung-directed autoreactivity, paint a strong picture of inflammatory milieu-directed responses.

An important question going forward will be the geographical origin of these autoreactive breaks. Indeed, while germinal centers have been identified as absent in the draining lymph node, it will be important to identify if the EF response in secondary lymph nodes is sufficient to drive these relaxed-tolerance responses. In this scenario, inflammatory lung tissue would be sufficiently generated in severe disease to result in drainage of lung-based antigen to local draining lymph nodes resulting in responses against a mix of lung antigen, viral antigen, and activated immune components. An alternative scenario would place the generation of these responses within the lung itself. In lupus nephritis, engraftment of plasma cells into the kidney parenchyma is an excellent predictor of disease severity, and it is not yet clear to what extent those cells develop locally. This scenario, while possible, seems less likely as it would require the recirculation of these cells from the local tissue out into the blood where they could be assessed in our PBMC-based study.

Regardless of the origin of these responses, recent work by Grant et. al. finds that a significant fraction (7.5%) of the total cellularity obtained in bronchoalveolar lavage fluid within 48h of intubation were plasma cells. Deeper analysis of those data suggest that a substantial fraction of those cells can be identified expressing the *IGHG1* transcript. While a more direct analysis of clonotype sharing is needed, it seems highly likely that the early emergence of these cells in the blood are linked to lung plasma cell engraftment. This would suggest that these breaks in peripheral tolerance, and subsequent emergence of lung-targeted autoreactivity are acting both locally and systemically with significant implications in tissue pathology and disease course. It will be important to understand the persistence of these cells in local lung tissue and understand their relevance to ongoing symptomology in recovered patients with persisting pulmonary effects.

While the evidence presented here for relaxed peripheral tolerance is clear, an important consideration of these findings is that this autoreactive profile does not extend to *all* antigens present in the local environment. Despite broad autoreactive profiles in these patients, they did not break tolerance against some antigens commonly found in autoimmune diseases such as anti-dsDNA. In addition to the inflammatory destruction of lung tissue present in severe disease, neutrophil activation and NET formation has been confirmed in severe COVID-19 making exposure of these developing autoreactive responses to DNA a near certainty. This was also true of ‘reflex’ ANA antigens such as La, Sm, and Ro – while ANAs were generally positive in these patients, the autoantigens more specific to SLE remained negative during acute infection. Although several explanations are possible, the most intriguing is that B cell responses against these antigens are sufficiently selected against during central tolerance to avoid peripherally derived autoreactivities. If this is indeed the case, the autoreactive profiles we are observing in these patients may be specific to breaks in peripheral tolerance. In this case, anti-carbamylated protein responses previously observed in correlation with severe autoimmunity^29^ and associated with NET formation^30^ would be a biomarker of the acute relaxation of peripheral tolerance, rather than a generalized marker of chronic progressive disease. As a readily available clinical test, it could also serve as an indication for more aggressive B-cell based therapy in active infection.

Perhaps the most important finding in this study in relation to patient health is the observed retraction of this autoreactive compartment through both serological, and repertoire-based follow up. An unexpected finding was the complete loss of highly expanded antiviral ASC clonotypes at acute timepoints from the memory repertoire – even those that ‘correctly’ identified viral targets with reasonably high affinity. As other, non-IgG1-based compartments showed greater connectivity to the recovered repertoire, this suggests that EF vs. GC responses in these patients are strictly segregated. These data would line up with previous findings in mice suggesting short-lived plasmablasts are the main output of the EF response, and suggests that clonotypes selected for EF responses are not recovered or reincorporated into the germinal center environment. It also suggests that autoreactivity following recovery could simply be a remnant of the acute response, with no basis in the underlying immune memory. It is important to point out, however, that not all autoreactivity returned to baseline in these patients over 6 months, and it is possible that additional niches capable of supporting autoreactive memory exist not addressed in our current methodology.

This lingering-autoreactivity model has critical implications in long-term recovery. We and others have repeatedly demonstrated broad autoreactivity in class-switched ASC compartments directly linked to disease pathology through immune component and lung-tissue directed autoreactivity. While the repertoire underpinning these responses did not appear to persist into the memory compartment in our study cohort, there is no reason to believe that these antibodies wouldn’t persist for the ‘normal’ half-life of circulating IgG1 antibodies. In this case, patients who experience autoreactive breaks in acute infection have received the equivalent of a high-dose infusion of generally autoreactive antibodies mixed with their antiviral response. It is highly possible, if not likely, that patients who recover from these acute tolerance breaks would continue to feel the effects of these antibodies, with a variety of manifestations in accordance with specific autoantigen targeting. <u>In this scenario, cycling the patient’s IgG fraction could have a large impact on resolving symptoms</u>. Indeed, this model is supported by anecdotal reports of resolving long-hauler symptoms following vaccination, whereby simple IgG cycling to viral-directed antibodies would reduce the fraction of autoreactive antibodies in the blood. If this is the case, IVIG infusion or plasma exchange therapy may have similar utility and warrants urgent clinical investigation in patients with severe disease at early timepoints. Notably, both therapeutic plasma exchange^31^ and IVIG^32^ have already shown positive results in small case series but need to be examined in larger, multicenter, randomized clinical trials.

Altogether, this study contextualizes the broad autoreactive profiles now well-established in severe COVID-19 as a functional consequence of a loss of selection stringency within the emerging ASC compartment early on in infection. These responses are contemporaneous with antiviral responses, and indeed, some antiviral clonotypes are bi-specific for autoantigens. The results of this low-selection environment are broad breaks in peripheral tolerance, focused specifically on the local viral/immune/lung inflammatory milieu. These breaks can have pathological consequences, and efforts should be made to understand if strengthened B cell interventions, alongside plasma exchange therapy or IVIG infusions could stand to benefit patients with acute severe disease. Anti-carbamylated protein responses have emerged as a clinically available biomarker for breaks in peripheral tolerance and could be easily incorporated into routine patient testing to serve as an indication for these stronger immune-targeted therapies. Finally, we document the resolution of these responses, the reestablishment of normal selection stringency into the recovered ASC compartment, and the loss of generalized autoreactivity in broad clinical testing in patients with significant autoimmune manifestations.

## Methods

### Human participants

All research was approved by the Emory University Institutional Review Board (Emory IRB nos. IRB00058507, IRB00057983 and IRB00058271) and was performed in accordance with all relevant guidelines and regulations. Written informed consent was obtained from all participants or, if they were unable to provide informed consent, from designated healthcare surrogates. Healthy individuals (n = 20) were recruited using promotional materials approved by the Emory University Institutional Review Board. Individuals with COVID-19 (n = 19) were recruited from Emory University Hospital, Emory University Hospital Midtown and Emory St. Joseph’s Hospital (all Atlanta, USA). All non-healthy individuals were diagnosed with COVID-19 by PCR amplification of SARS-CoV-2 viral RNA obtained from nasopharyngeal or oropharyngeal swabs. Individuals with COVID-19 were included in the study if they were between 18 to 80 years of age, were not immunocompromised and had not been given oral or intravenous corticosteroids within the preceding 14 d. Peripheral blood was collected in either heparin sodium tubes (PBMCs) or serum tubes (serum; both BD Diagnostic Systems). Baseline individual demographics are included in Supplementary Table 1. Study data were collected and managed using REDCap electronic data capture tools hosted at Emory University.

### Peripheral blood mononuclear cell isolation and plasma collection

Peripheral blood samples were collected in heparin sodium tubes and processed within 6 h of collection. PBMCs were isolated by density gradient centrifugation at 1,000g for 10 min. Aliquots from the plasma layer were collected and stored at −80 °C until use. PBMCs were washed twice with RPMI at 500g for 5 min. Viability was assessed using trypan blue exclusion, and live cells were counted using an automated hemocytometer.

### Flow cytometry

Isolated PBMCs (2 × 106) were centrifuged and resuspended in 75 μl FACS buffer (PBS + 2% FBS) and 5 μl Fc receptor block (BioLegend, no. 422302) for 5 min at room temperature. For samples stained with anti-IgG, it was observed that Fc block inappropriately interfered with staining, so a preincubation step of the anti-IgG alone for 5 min at 22 °C was added before the addition of the block. Next, 25 μl of antibody cocktail (Supplementary Table 3) was added (100 μl staining reaction), and samples were incubated for 20 min at 4 °C. Cells were washed in PBS, and resuspended in a PBS dilution of Zombie NIR fixable viability dye (BioLegend, no. 423106). Cells were washed and fixed at 0.8% paraformaldehyde (PFA) for 10 min at 22 °C in the dark before a final wash and resuspension for analysis.

Cells were analyzed on a Cytek Aurora flow cytometer using Cytek SpectroFlo software. Up to 3 × 106 cells were analyzed using FlowJo v10 (Treestar) software.

### Analysis software

Computational analysis was carried out in R (v3.6.2; release 12 Dec 2019). Heat maps were generated using the pheatmap library (v1.0.12), with data pre-normalized (log-transformed z-scores calculated per feature) before plotting. Custom plotting, such as mutation frequency violin plots, was performed using the ggplot2 library for base analysis, and then post-processed in Adobe Illustrator. Alluvial plotting was performed using the ggalluvial package with post-processing in Adobe Illustrator. Clonotype connectivity analysis was carried out using the R-based ‘vegan’ package, and then visualized through ‘pheatmap’ before post-processing in Adobe Illustrator. Statistical analyses were performed directly in R, or in GraphPad Prism (v8.2.1).

Analyses on the single cell VDJ annotated sequences were performed using the Immcantation tool suite (http://www.immcantation.org) version 4.1.0 pipeline in Docker. This suite contains SHazaM for statistical analysis of somatic hypermutation (SHM) patterns as described in (Gupta et al., 2015), and BASELINe (Bayesian estimation of Antigen-driven SELectIoN) for analysis of selection pressure as described in (Yaari et al., 2012). Visualizations were generated in R using the SHazaM package (version 1.0.2) and then post-processed in Adobe Illustrator.

### Flow cytometry and sorting of B cell subsets for repertoire sequencing

Frozen cell suspensions were thawed at 37 °C in RPMI + 10% FCS and then washed and resuspended in FACS buffer (PBS + 2% FCS). The cells were incubated with a mix of fluorophore-conjugated antibodies for 30 min on ice. The cells were washed in PBS and then incubated with the live/dead fixable aqua dead cell stain (Thermo Fisher) for 10 min at 22 °C. After a final wash in FACS buffer, the cells were resuspended in FACS buffer at 107 cells per ml for cell sorting on a three-laser BD FACS (BD Biosciences).

For single-cell analysis, total ASCs were gated as CD3−CD14−CD16−CD19+CD38+CD27+ single live cells, whereas naive B cells were gated as CD3−CD14−CD16−CD19+CD27−IgD+CD38+ single live cells.

For bulk sequencing preparations, B cells were enriched using StemCell’s Human Pan-B Cell Enrichment Kit (no. 19554; negative selection of CD2, CD3, CD14, CD16, CD36, CD42b, CD56, CD66b and CD123). CD138+ ASCs were enriched further using CD138+ selection beads according to the manufacturer’s instructions (Miltenyi Biotec, no. 130-051-301).

### Single cell V(D)J repertoire library preparation and sequencing

Cells were counted immediately using a hemocytometer and adjusted to 1,000 cells per μl to capture 10,000 single cells per sample loaded in the 10× Genomics Chromium device according to the manufacturer’s standard protocol (Chromium Next GEM Single Cell V(D)J Reagent Kits, v1.1). The 10× Genomics v2 libraries were prepared using the 10x Genomics Chromium Single Cell 5′ Library Construction Kit per the manufacturer’s instructions. Libraries were sequenced on an Illumina NovaSeq (paired-end; 2 × 150 bp; read 1:26 cycles; i7 index: 8 cycles, i5 index: 0 cycles; read 2: 98 cycles) such that more than 70% saturation could be achieved with a sequence depth of 5,000 reads per cell.

### Carbodiimide coupling of microspheres to SARS-CoV-2 antigens

Two SARS-CoV-2 proteins were coupled to MagPlex Microspheres of different regions (Luminex). Nucleocapsid (N) protein expressed from Escherichia coli (N-terminal His6) was obtained from Raybiotech (230-01104-100) and the RBD of spike (S) protein expressed from HEK293 cells was obtained from the laboratory of J. Wrammert63 at Emory University. Coupling was carried out at 22 °C following standard carbodiimide coupling procedures. Concentrations of coupled microspheres were confirmed by Bio-Rad T20 Cell Counter.

### Luminex proteomic assays for measurement of anti-antigen antibody

Approximately 50 μl of coupled microsphere mix was added to each well of 96-well clear-bottom black polystyrene microplates (Greiner Bio-One) at a concentration of 1,000 microspheres per region per well. All wash steps and dilutions were accomplished using 1% BSA, 1× PBS assay buffer. Sera were assayed at 1:500 dilutions and surveyed for antibodies against N or RBD. After a 1-h incubation in the dark on a plate shaker at 800 r.p.m., wells were washed five times in 100 μl of assay buffer, using a BioTek 405 TS plate washer, then applied with 3 μg ml−1 PE-conjugated goat anti-human IgA, IgG and/or IgM (Southern Biotech). After 30 min of incubation at 800 r.p.m. in the dark, wells were washed three times in 100 μl of assay buffer, resuspended in 100 μl of assay buffer and analyzed using a Luminex FLEXMAP 3D instrument (Luminex) running xPONENT 4.3 software. MFI using combined or individual detection antibodies (anti-IgA, anti-IgG or anti-IgM) was measured using the Luminex xPONENT software. The background value of assay buffer was subtracted from each serum sample result to obtain MFI minus background (MFI-B; net MFI).

### Statistical analysis

Statistical analysis was carried out using Prism (GraphPad). For each experiment, the type of statistical testing, summary statistics and levels of significance can be found in the figures and corresponding legends. All measurements displayed were taken from distinct samples.

### High-Throughput Surface Plasmon Resonance

HT-SPR data was collected through single-cycle kinetic analysis against either SARS-CoV-2 nucleocapsid or spike trimer (S2P). Monoclonal antibodies were pre-screened for antigen binding through luminex-based multiplex binding assessment (above), and select antibodies were analyzed for binding affinity testing. All data was collected with 1:1 referencing collected in real time on a Nicoya Alto HT-SPR with 8 referenced channels running in parallel on carboxyl-coated sensors. Ligand binding/regeneration conditions for each antigen were as follows:

*S2P* – SARS-CoV-2 spike trimer was resuspended in tris acetate buffer, pH4.5, and immobilized onto an EDC/NHS-activated carboxyl sensor for 5 min at 50ug/ml. Regeneration of the sensor was performed using Glycine HCl, pH 2.5 for 1 min.

*Nucleocapsid* – SARS-CoV-2 nucleocapsid protein was resuspended in tris acetate buffer, pH6, and immobilized onto an EDC/NHS-activated carboxyl sensor for 5 min at 50ug/ml. Regeneration of the sensor was performed using Glycine HCl, pH 3 for 1 min.

All single-curve kinetics were performed with 5, 3-fold analyte dilutions with final concentrations between 222nM and 914pM. Analytes were run in PBST, with interactions collected at 25C.

### B cell binding assay

2-3 million healthy donor PBMCs were incubated with 5ug of mAb at 40C for 30 min. The cells were washed with 30x volume FACS buffer (1xPBS, 2%FBS) and subsequently stained with Ab to CD3, CD19, CD27, IgD and IgG, as well as with Zombie NIR. Staining was completed with 0.8 % Paraformaldehyde for fixation. Flow Cytometry analysis was performed on CytoFLEX (BD Biosciences). Dead cells and doublets were excluded. The median fluorescence intensity (MFI) of mAb (IgG) was determined on naïve B cell population.

### Monoclonal antibody selection and production

Monoclonal antibodies were selected for production from the single-cell repertoire data obtained from patient ICU-1. Individual cells were clustered into clonotypes, and then assessed for clonotype size, nucleotide mutation frequency, isotype, and connectivity between sorted populations. Through progressive filtering, clonotypes were selected that met the following criteria:

1. Contained at least one IgG1 member

2. Had at least one member with a mutation frequency of <1%

3. Had at least one member in the ASC compartment, the CD27-compartment, or contained members in both.

With those criteria met, all expanded clonotypes (>5 individual cells identified in the clonotype), and all *IGHV4-34*^+^ members were selected for monoclonal antibody production and screening – 55 clonotypes in all. The most frequently repeated BCR sequence from each clonotype was provided to Genscript for antibody production on a standard IgG1 backbone.

### Clinical autoreactivity testing

For autoimmune biomarker analysis frozen plasma was shipped on dry ice to Exagen, Inc. (Vista, California, USA) which has a clinical laboratory accredited by the College of American Pathologists (CAP) and certified under the Clinical Laboratory Improvement Amendments (CLIA). Thawed plasma was aliquoted and distributed for the following tests: anti-nuclear antibodies (ANA) were measured using enzyme-linked immunosorbent assays (ELISA) (QUANTA Lite; Inova Diagnostics) and indirect immunofluorescence (IFA) (NOVA Lite; Inova Diagnostics); anti-double-stranded DNA (dsDNA) antibodies were also measured by ELISA and were confirmed by IFA with Crithidia luciliae; extractable nuclear antigen autoantibodies (anti-Sm, anti-SS-B/La IgG, anti-Scl-70 IgG, anti-U1RNP IgG, anti-RNP70 IgG, anti-CENP IgG, anti-Jo-1 IgG, and anti-CCP IgG) as well as Rheumatoid Factor (RF) IgA and IgM were measured using the EliA test on the Phadia 250 platform (ThermoFisher Scientific); IgG, IgM, and IgA isotypes of anti-cardiolipin and anti-β2-glycoprotein, as well as anti-Ro52, anti-Ro60, anti-GBM, anti-PR3, and anti-MPO were measured using a chemiluminescence immunoassay (BIO-FLASH; Inova Diagnostics); anti-CarP, anti-RNA-pol-III, and the IgG and IgM isotypes of anti-PS/PT were measured by ELISA (QUANTA Lite; Inova Diagnostics), while C-and P-ANCA were measured by IFA (NOVA Lite; Inova Diagnostics). All assays were performed following the manufacturer’s instructions.

### CDI Arrays

For broad road plasma autoreactivity testing, 20ul frozen patient plasma was sent to CDI LABS for autoantigen testing on the HuProt platform. The chip-based array contains 21,000 human protein targets manufactured through yeast expression and printing on a nitrocellulose substrate. Plasma samples were diluted 1:1000 prior to incubation on the array, and the array was developed with an IgG-specific secondary fluorescent probe. Z-scores were calculated and reported for each protein target based on duplicate spot fluorescence and internal negative control fluorescence.

### BALF Plasma Cell Gene Expression

To assess the constant region gene expression in BALF-derived ASCs, data was retrospectively analyzed from the UCSC data browser available here: https://www.nupulmonary.org/covid-19-ms1. Briefly, these data are representative of 10 ICU patients whose BALF was collected within 48 hours of intubation, with total isolated cells sequenced using the 10x single cell transcriptomics platform. Patient information and full methods are available in the associated manuscript, Grant et. al. 2021.

### MENSA Generation

Medium enriched for newly synthesized antibodies (MENSA) was generated by isolating, washing, and culturing ASC-containing peripheral blood mono-nuclear cells (PBMC) from blood using a modified procedure previously described (REF). PBMC were isolated by centrifugation (1,000 ×g; 10 min) using Lymphocyte Separation Media (Corning) and Leucosep tubes (Greiner Bio-One). Five washes with RPMI-1640 (Corning) were performed to remove serum immunoglobulins (800 xg; 5 min), with erythrocyte lysis (3 mL; 3 min) after the second wash and cell counting after the fourth. Harvested PBMCs were cultured at 106 cells/mL in R10 Medium (RPMI-1640, 10% Sigma FBS, 1% Gibco Antibiotic/Anti-mycotic) on a 12-well, sterile, tissue culture plate for 24 h at 37° C and 5% CO2. After incubation, the cell suspension was centrifuged (800 ×g; 5 min) and the supernatant (MENSA) was separated from the PBMC pellet, aliquoted and stored at -80°C for testing.

### COVID-19 Multiplex Immunoassay

SARS-CoV-2 antigens were coupled to MagPlex Microspheres of spectrally distinct regions via carbodiimide coupling and tested against patient samples as previously described (2). Results were analyzed on a Luminex FLEXMAP 3D instrument running xPonent 4.3 software. Median fluorescent intensity (MFI) using combined or individual PE-conjugated detection antibodies (anti-IgA/anti-IgG/anti-IgM) was measured using the Luminex xPONENT software on Enhanced PMT setting. The background value of assay buffer or R10 media was subtracted from the serum/plasma or MENSA results, respectively, to obtain MFI minus background (net MFI). Serum and plasma samples were tested at 1:500 dilution and MENSA was tested undiluted.

### Selection of Antigens

#### MENSA and Serum samples

Four recombinant SARS-CoV-2 Ags were used in this study. The N protein (catalog no. Z03480; expressed in Escherichia coli), the S1 domain (aa 16–685; catalog no. Z03485; expressed in HEK293 cells) of the spike protein, and the S1-RBD (catalog no. Z03483; expressed in HEK293 cells) were purchased from GenScript. The S1-NTD (aa 16–318) was custom synthesized by GenScript. Each protein was expressed with an N-terminal His6-tag to facilitate purification, >85% pure and appeared as a predominant single band on SDS-PAGE analysis.

#### Monoclonal Antibody testing

RBD (catalog no. Z03483; expressed in HEK293 cells) and Nucleocapsid protein (catalog no. Z03480; expressed in Escherichia coli), were purchased from GenScript (same as the first version). S1 (catalog no. S1N-C52H3; HEK293), S2 (catalog no. S2N-C52H5; HEK293) and S1 N-terminal domain (NTD; catalog no. S1D-C52H6; HEK293) were purchased from ACROBiosystems. The C-terminus sequence of ORF3a (Accession: QHD43417.1, amino acids 134-275 plus N-terminal His6-Tag) was sent to Genscript for custom protein expression in E. coli.

## Supporting information

Main tables

## Data Availability

Relevant data will be made available as necessary following peer-review

## Acknowledgments

We would like to thank S. Auld, W. Bender, L. Daniels, B. Staitieh, C. Swenson, and A. Truong for their expertise and support of our research. We would also like to thank the nurses, staff, and providers in the 71 ICU in Emory University Hospital Midtown and the 2E ICU in Emory Saint Joseph’s Hospital. We would like to thank S. Rey, S. Demers, M. Hammons, A. Sace, and R. LaFon, and the Sanz/Lee sample processing teams for aid in sample preparation and serological screening. Finally, we would acknowledge Dr. Luisa Morales-Nebreda for her guidance in use of previously published datasets.

---

**Supplemental Figure 1.**
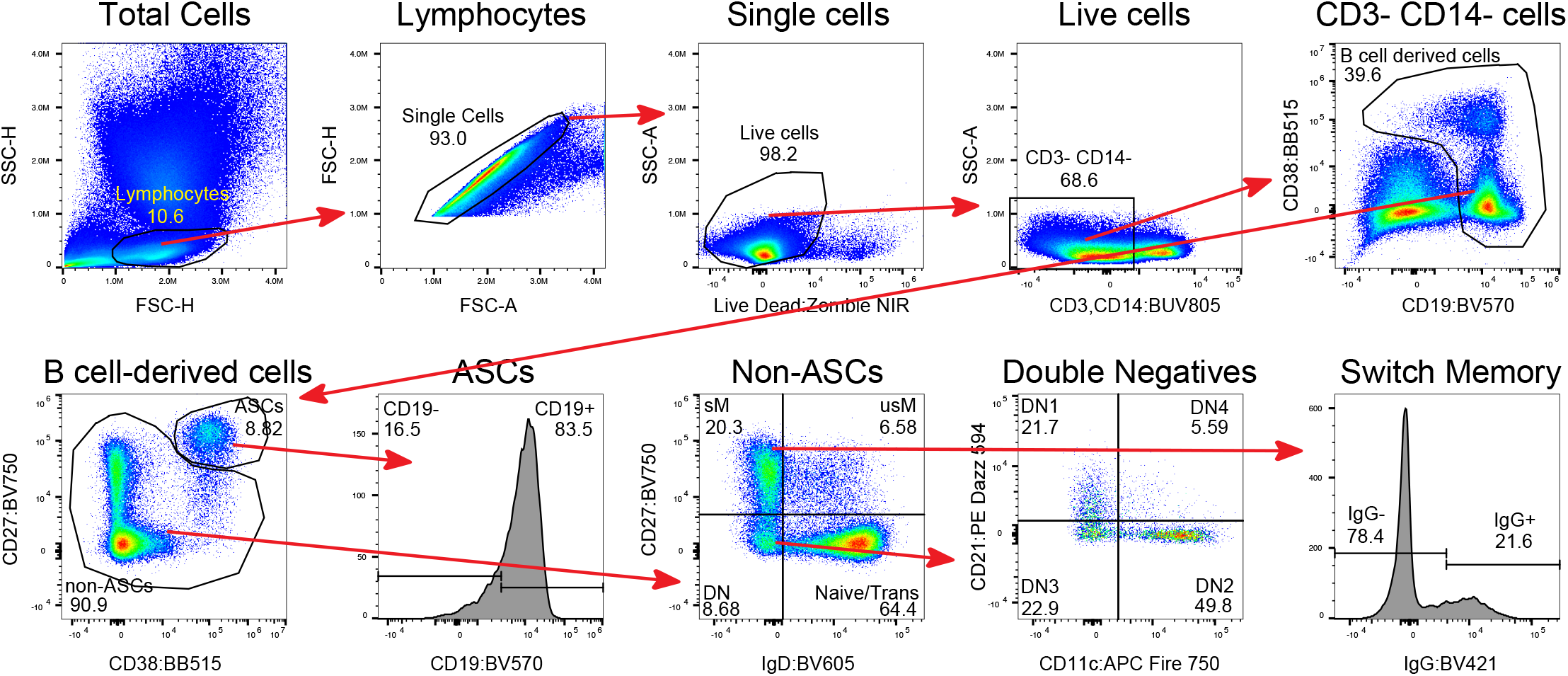
Gating strategy for high-dimension flow cytometry. Progressive gating strategy for high-dimensional flow cytometry. Label above plot indicated pre-gating population from previous plot.

**Supplemental Figure 2.**
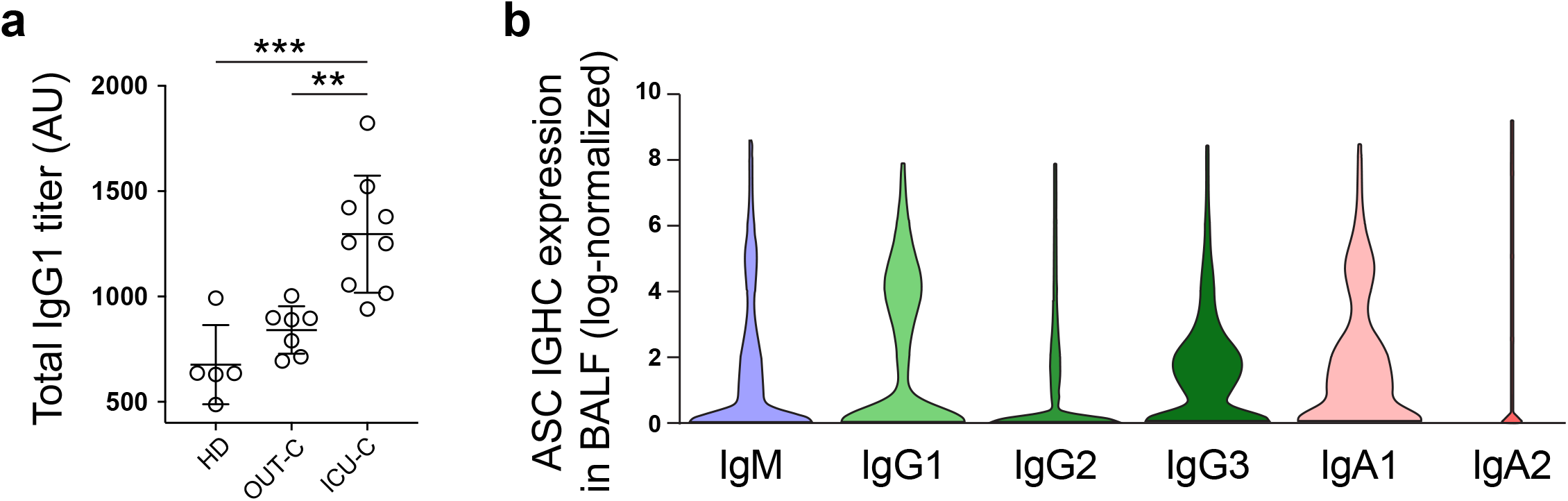
IgG1 class switching in the plasma and lung. (a) Bulk IgG1 assessment in HD, OUT-C, or ICU-C cohorts. (b) Gene expression of indicated constant region in ASCs identified in the bronchoalveolar fluid from 10 ICU patients. Retrospective analysis of data collected by Grant et. al.

**Supplemental Figure 3.**
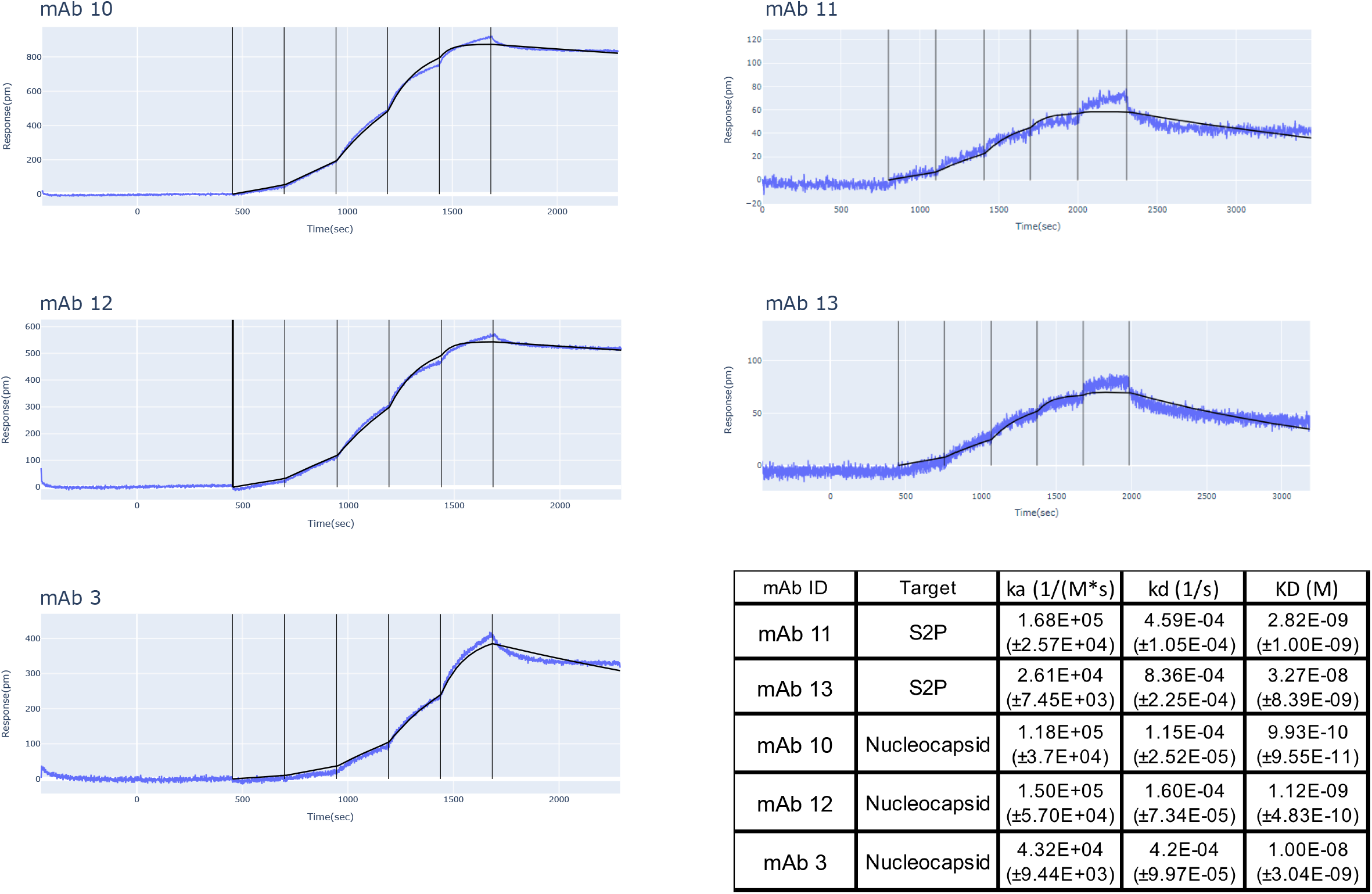
Data fitting for mAb affinity testing via HT-SPR. Representative raw data (blue), and model fitting (black) are displayed for each of the 5 antibodies tested for affinity via HT-SPR. Summary table displays the target, on rate (Ka), off rate (Kd), and affinity (KD), with associated standard deviations in parentheses.

**Supplemental Figure 4.**
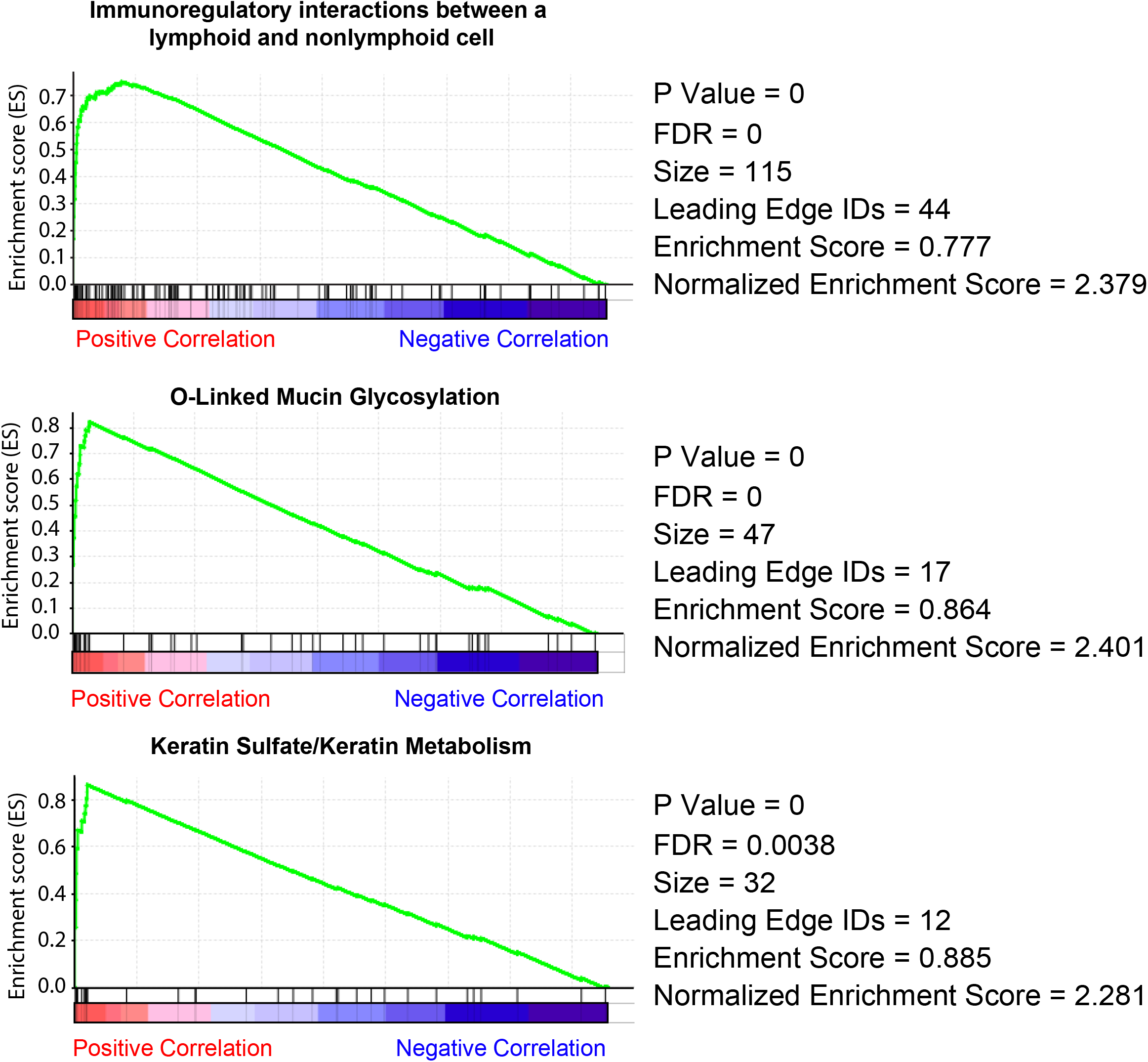
GSEA analysis of Reactome pathways. GSEA analysis of CDI autoantigen array results from patient ICU-1 plasma. Top 3 enriched pathways (ranked on p value) are shown.

## References

1. Andersen, K.G., Rambaut, A., Lipkin, W.I., Holmes, E.C. & Garry, R.F. The proximal origin of SARS-CoV-2. Nat Med 26, 450–452 (2020).

2. Chen, X. et al. Detectable Serum Severe Acute Respiratory Syndrome Coronavirus 2 Viral Load (RNAemia) Is Closely Correlated With Drastically Elevated Interleukin 6 Level in Critically Ill Patients With Coronavirus Disease 2019. Clin Infect Dis 71, 1937–1942 (2020).

3. Henderson, L.A. et al. On the Alert for Cytokine Storm: Immunopathology in COVID-19. Arthritis Rheumatol 72, 1059–1063 (2020).

4. Group, R.C. et al. Dexamethasone in Hospitalized Patients with Covid-19. N Engl J Med 384, 693–704 (2021).

5. Cao, X. COVID-19: immunopathology and its implications for therapy. Nat Rev Immunol 20, 269–270 (2020).

6. Mathew, D. et al. Deep immune profiling of COVID-19 patients reveals distinct immunotypes with therapeutic implications. Science 369 (2020).

7. Kaneko, N. et al. Loss of Bcl-6-Expressing T Follicular Helper Cells and Germinal Centers in COVID-19. Cell 183, 143–157 e113 (2020).

8. Woodruff, M.C. et al. Extrafollicular B cell responses correlate with neutralizing antibodies and morbidity in COVID-19. Nat Immunol 21, 1506–1516 (2020).

9. Hoehn, K.B. et al. Cutting Edge: Distinct B Cell Repertoires Characterize Patients with Mild and Severe COVID-19. J Immunol (2021).

10. Sosa-Hernandez, V.A. et al. B Cell Subsets as Severity-Associated Signatures in COVID-19 Patients. Front Immunol 11, 611004 (2020).

11. Nielsen, S.C.A. et al. Human B Cell Clonal Expansion and Convergent Antibody Responses to SARS-CoV-2. Cell Host Microbe 28, 516–525 e515 (2020).

12. Jenks, S.A. et al. Distinct Effector B Cells Induced by Unregulated Toll-like Receptor 7 Contribute to Pathogenic Responses in Systemic Lupus Erythematosus. Immunity 49, 725–739 e726 (2018).

13. Jenks, S.A., Cashman, K.S., Woodruff, M.C., Lee, F.E. & Sanz, I. Extrafollicular responses in humans and SLE. Immunol Rev 288, 136–148 (2019).

14. Tipton, C.M. et al. Diversity, cellular origin and autoreactivity of antibody-secreting cell population expansions in acute systemic lupus erythematosus. Nat Immunol 16, 755–765 (2015).

15. Zhang, Y. et al. Coagulopathy and Antiphospholipid Antibodies in Patients with Covid-19. N Engl J Med 382, e38 (2020).

16. Bastard, P. et al. Autoantibodies against type I IFNs in patients with life-threatening COVID-19. Science 370 (2020).

17. Gagiannis, D. et al. Clinical, Serological, and Histopathological Similarities Between Severe COVID-19 and Acute Exacerbation of Connective Tissue Disease-Associated Interstitial Lung Disease (CTD-ILD). Front Immunol 11, 587517 (2020).

18. Bowles, L. et al. Lupus Anticoagulant and Abnormal Coagulation Tests in Patients with Covid-19. N Engl J Med 383, 288–290 (2020).

19. Wang, E.Y. et al. Diverse functional autoantibodies in patients with COVID-19. Nature (2021).

20. Haddad, N.S. et al. Novel immunoassay for diagnosis of ongoing Clostridioides difficile infections using serum and medium enriched for newly synthesized antibodies (MENSA). J Immunol Methods 492, 112932 (2021).

21. Grant, R.A. et al. Circuits between infected macrophages and T cells in SARS-CoV-2 pneumonia. Nature 590, 635–641 (2021).

22. Yaari, G., Uduman, M. & Kleinstein, S.H. Quantifying selection in high-throughput Immunoglobulin sequencing data sets. Nucleic Acids Res 40, e134 (2012).

23. Reed, J.H., Jackson, J., Christ, D. & Goodnow, C.C. Clonal redemption of autoantibodies by somatic hypermutation away from self-reactivity during human immunization. J Exp Med 213, 1255–1265 (2016).

24. Haddad, N.S. et al. One-Stop Serum Assay Identifies COVID-19 Disease Severity and Vaccination Responses. Immunohorizons 5, 322–335 (2021).

25. Premkumar, L. et al. The receptor binding domain of the viral spike protein is an immunodominant and highly specific target of antibodies in SARS-CoV-2 patients. Sci Immunol 5 (2020).

26. McAdoo, S.P. & Pusey, C.D. Anti-Glomerular Basement Membrane Disease. Clin J Am Soc Nephrol 12, 1162–1172 (2017).

27. Satoh, M. et al. Prevalence and sociodemographic correlates of antinuclear antibodies in the United States. Arthritis Rheum 64, 2319–2327 (2012).

28. Luo, X. et al. Prognostic Value of C-Reactive Protein in Patients With Coronavirus 2019. Clin Infect Dis 71, 2174–2179 (2020).

29. Ceccarelli, F. et al. Anti-carbamylated protein antibodies as a new biomarker of erosive joint damage in systemic lupus erythematosus. Arthritis Res Ther 20, 126 (2018).

30. Clarke, J. NETs revealed as source of carbamylated proteins in RA. Nat Rev Rheumatol 17, 4 (2021).

31. Truong, A.D. et al. Therapeutic plasma exchange for COVID-19-associated hyperviscosity. Transfusion 61, 1029–1034 (2021).

32. Tzilas, V., Manali, E., Papiris, S. & Bouros, D. Intravenous Immunoglobulin for the Treatment of COVID-19: A Promising Tool. Respiration 99, 1087–1089 (2020).

